# Pareto-based evaluation of national responses to COVID-19 pandemic shows that saving lives and protecting economy are non-trade-off objectives

**DOI:** 10.1101/2020.06.27.20141747

**Authors:** Marek Kochańczyk, Tomasz Lipniacki

## Abstract

Countries worldwide have adopted various strategies to minimize the socio-economic impact of the ongoing COVID-19 pandemic. Stringency of imposed measures universally reflects the standpoint from which protecting public health and saving economy are seen as contradictory objectives. We analyzed epidemic trajectories of 25 highly developed countries and 10 US states in the mobility reduction *vs*. reproduction number plane to show that the consequence of delayed lockdown is a surge of infections and deaths, that is ultimately suppressed by stringent and long-lasting quarantine. As a consequence, cumulative mobility reduction and population-normalized cumulative number of COVID-19-associated deaths are significantly correlated and this correlation increases with time. We thus demonstrated that, as long as epidemic suppression is the aim, the trade-off between the death toll and economic loss is illusory: high death toll correlates with deep and long-lasting lockdown and severe economic downturn.

## Introduction

The COVID-19 epidemic is a major threat to both public health and global economy. To minimize death toll and economic loss, countries worldwide have assumed various strategies (not always officially declared). Two main strategies to counteract the epidemic spread are eradication and suppression. Countries such as China, South Korea, Japan, Taiwan, New Zealand, Australia, and others, decided to fully eradicate the disease on their territories and then limit and control incoming travelers. Geographic isolation helps to implement such strategy. Most European countries have chosen the suppression strategy that aims to limit the exponential growth of the outbreak and then reduce daily new cases to a small number. Both the eradication and the suppression strategy are founded on the expectation that a vaccine will be developed in near future, and that massive vaccination will ultimately enable governments to lift imposed restrictions. A third strategy, in which it is anticipated that, on the contrary, no effective vaccine will be available soon and thus long-term containment is not realistic, is to rely on mitigation and ultimately acquisition of herd immunity. This strategy had been initially declared in the UK but was promptly abandoned due to a rapid increase in the death toll and the threat of overwhelming the health system.

In the conceptual framework of mathematical models of infectious diseases, any policy that aims at reducing transmission of the virus seeks to decrease the effective reproduction number. Within S(E)IR-type epidemic models [1], the reproduction number *R* is equal to the ratio of the contact rate *β* of the infectious and the susceptible individuals and the removal rate *γ* of the infectious individuals, *R* = *β*/*γ*. In terms of policy-making, the contact rate *β* can be reduced by prohibiting mass gatherings (that increase the risk of super-spreading events), social distancing, and hygienic precautions. These countermeasures decrease the risk that a given contact is infectious. Reduction of is *β* may be also achieved through quarantine that obligates individuals to stay home or at least significantly limit their out-of-home activity. The removal rate *γ* can be increased by enhanced testing and contact tracing, which both enable prompt isolation of (potentially) infectious individuals. While a nation-wide quarantine (lockdown) is an effective mean of reducing *β* (and *R*) and saving lives, it hampers economic activity and may thus incur a significant loss of country GDP, in addition to psychological and social costs. Policy-making is thus intuitively understood as a balancing act between health and fiscal risks of the pandemic.

We show that when the strategies of multiple countries are juxtaposed, there is effectively no trade-off between the death toll and the economic loss. As the total number of deaths and cumulative lockdown are the two quantities that have to be jointly minimized, we use Pareto fronts to evaluate the European and North American suppression policies as well as East Asian eradication policies. We demonstrate that there is a strong correlation between the population-normalized cumulative number of COVID-19-assigned deaths and cumulative lockdown. A similar relation is observed for excess deaths and decrease of GDP in the first half of 2020. We interpret this result by dissecting the impact of testing and nation-wide quarantine on the effective reproduction number.

## Results

Severity of national lockdowns can be inferred from data on the reduction of mobility gathered in Google COVID-19 Community Mobility Reports [2]. We expect that the relative reduction of mobility in locations belonging to two categories—workplaces and places related to retail and recreation—is strongly associated with economic slowdown. We thus assume that the cumulative average mobility reduction in these two categories serves as a good indicator of the impact the epidemic has on economy.

In Fig. 1 we analyzed Western, Central, and South European countries with populations of at least five million, ten most populous US states, and Canada—that nearly unanimously adopted the suppression strategy—as well as Taiwan, South Korea and Japan—that adopted the eradication strategy. In all considered countries, human development index, that factors in life expectancy and standard of living, exceeds 0.8 [3]. Also, all considered locations are in the Northern Hemisphere, which minimizes the influence of seasonal effects related to weather and holidays [4]. We marked positions of the considered locations on the plane of cumulative mobility reduction (lockdown) *vs*. square root of the population-normalized cumulative number of COVID-19-assigned deaths as of July 1 (Fig. 1a) and November 1 (Fig. 1b) 2020.

**Figure 1:**
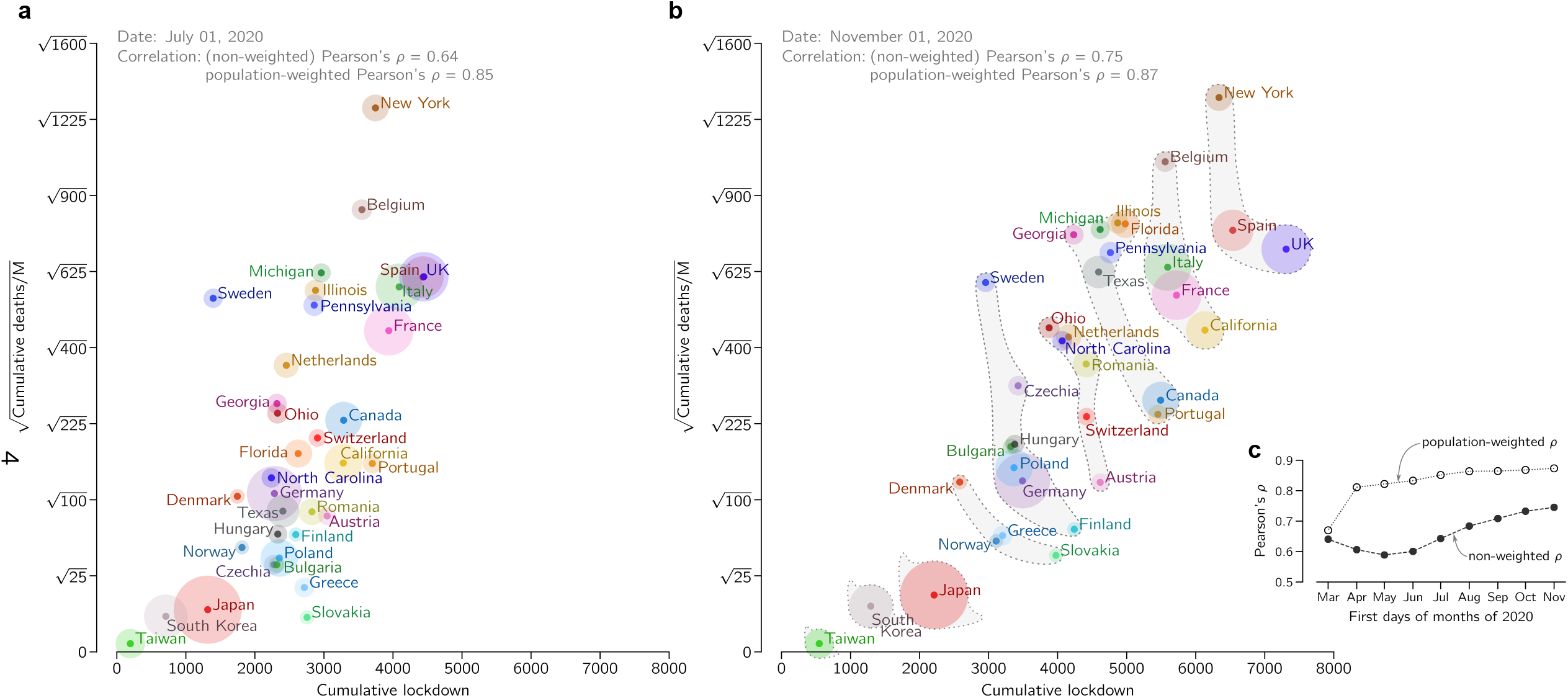
Cumulative mobility reduction (lockdown) and square root of the cumulative COVID-19-assigned deaths per million population for European and East Asian countries, ten US states, and Canada. (**a**) Positions of considered locations as of July 1, 2020. (**b**) Positions of considered locations as of November 1, 2020, and fuzzy Pareto fronts. In (a) and (b), disk area of each location is proportional to its population. (**c**) Pearson’s correlation coefficients between cumulative mobility reduction and square root of the population-normalized cumulative number of COVID-19-assigned deaths.

Figures 1a and 1b suggest high correlation between the population-normalized cumulative number of COVID-19-assigned deaths and the cumulative mobility reduction. Both these data series are normally distributed (whereas the set of non-transformed population-normalized cumulative numbers of COVID-19-assigned deaths does not satisfy the normality condition, see Methods for details). This allowed us to quantify interdependence of these two data series using Pearson’s correlation coefficient. For November 1, 2020, we obtained *ρ* = 0.75 and substantially higher population-weighted correlation coefficient, *ρ*_w_ = 0.87. Importantly, as demonstrated in Fig. 1b, both correlation coefficients, *ρ* and *ρ*_w_, increased in time. In Supplementary Figure S1 we show how positions of considered locations changed in time in the coordinate system of Fig. 1a and Fig. 1b.

The observed high correlation clearly indicates for a lack of trade-off between life/health cost and lockdown-induced economic/social loss. Consequently, when locations represented by such diagonally scattered data points are evaluated from the perspective of Pareto optimality, layered Pareto fronts are rather short (Fig. 1b). The trade-off between the considered objectives would be manifested by anti-correlation and correspondingly long Pareto fronts. In the Pareto approach, both objectives are considered simultaneously without assigning weights to them. By definition, each location from a considered layer performs worse (considering both criteria) than at least one location from the previous layer. We decided to group locations using “fuzzy” fronts to reduce the number of fronts and make them more robust with respect to small (and random) differences in values of considered objectives.

The first optimal front (*first* layer) consists of a single country, Taiwan, which has simultaneously the smallest cumulative lockdown and smallest population-normalized number of deaths; both next two optimal fronts also contain a solitary East Asian country, South Korea and Japan; the fourth optimal front contains four European countries: Slovakia, Norway, Greece, and Denmark. The first pessimal front (*last* layer) contains New York State (with the highest population-normalized number of deaths), Spain, and UK (with the highest cumulative lockdown); the second pessimal front is spanned by the state of California, and France, Italy and Belgium; the third, broad pessimal front groups several US states, Canada, and Portugal.

High correlation between cumulative lockdown and population-normalized cumulative number of deaths can be explained by analysis of trajectories of individual countries in the mobility reduction *vs*. effective reproduction number plane, see Fig. 2. The effective reproduction number *R* (equal *β*/*γ* in the SEIR model that we used to estimate *R* [5]) is calculated from the number of daily new registered cases as explained in Methods. In the countries that are close to the current Pareto-pessimal front (last layer), the epidemic had three distinguishable phases, depicted schematically in Fig. 3. In the initial phase, characterized by a high *R* value and no mobility reduction, *R* decreases due to suppression of potential super-spreading events and social distancing. In the next phase, *R* drops below 1 due to the reduction of mobility, which leads to a decrease of the contact rate *β*. When analyzing individual trajectories plotted in Fig. 2 we may notice that in the second phase the number of tests per case (indicated by line width and color intensity) remains constant or decreases (possibly due to saturation of the capacity of facilities that perform tests), suggesting that in this phase the removal rate *γ* remains constant or decreases. This confirms that the reduction of mobility, and thus *β*, was instrumental in bringing *R* below 1.

**Figure 2:**
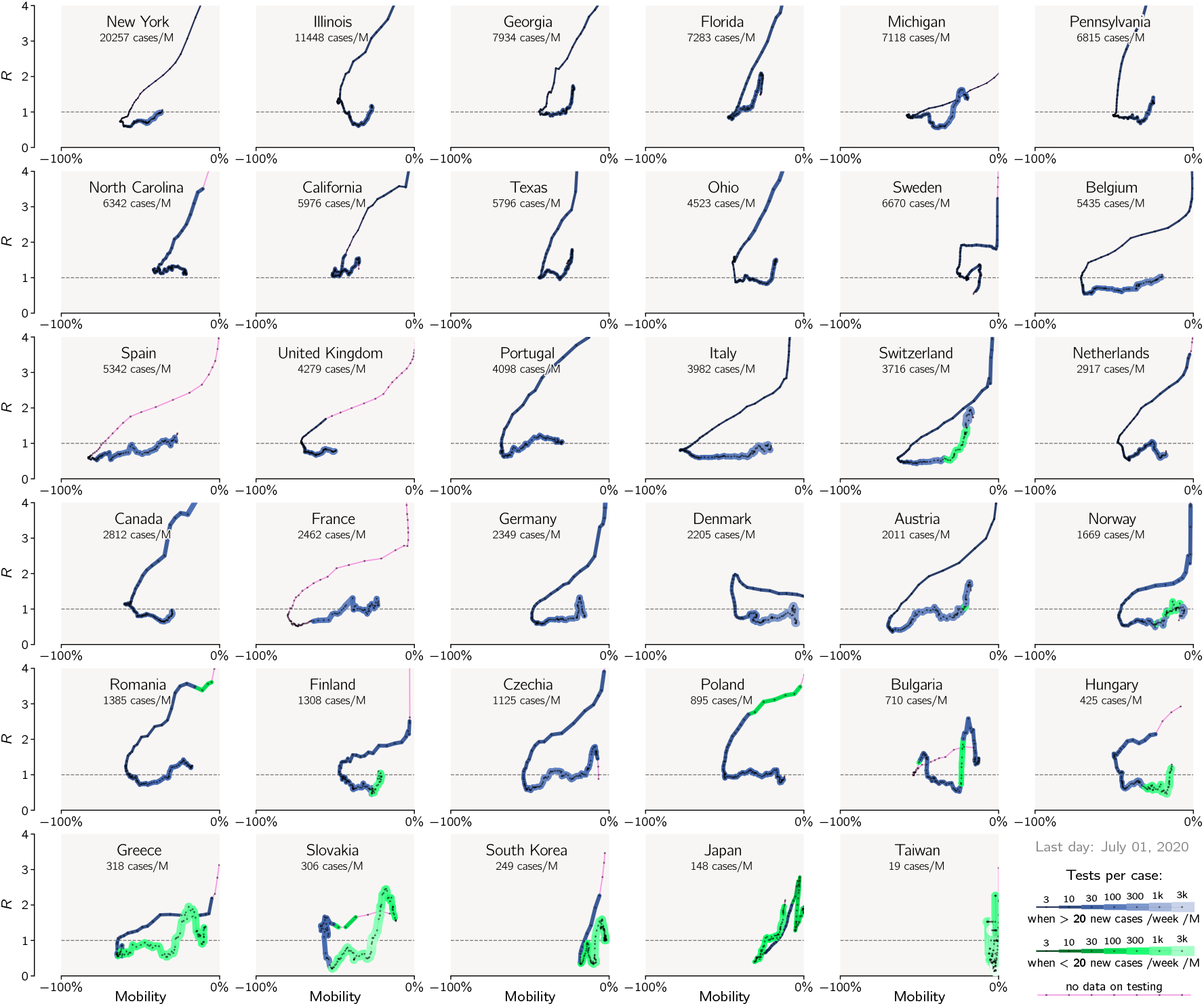
Epidemic trajectories of countries and states analyzed in Fig. 1 in the mobility reduction *vs*. effective reproduction number (*R*) plane. The value of *R* has been estimated based on the reported number of daily new cases. The trajectories begin in February and end on July 1, 2020, and have a single-day resolution. Line thickness increases with the reported number of tests per case (positive test).

**Figure 3:**
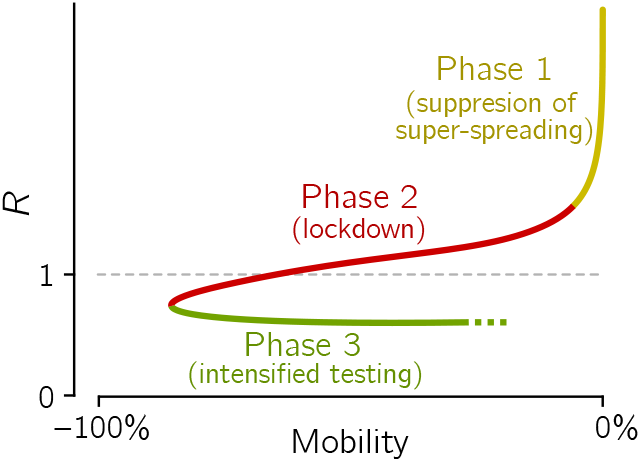
Scheme of a three-phase epidemic trajectory of the first wave typical for the most affected European countries that implemented effective lockdowns.

We digress here to note that the daily number of performed tests can be analyzed in relation to either the population size or the number of positive cases (positive tests). We think that this latter scaling is more adequate as it reflects the idea of contact tracing, when the individuals that had contact with an infected individual undergo testing. The bigger is the tested cluster of individuals, the higher is the probability that majority of the secondary infectious will be identified and “removed”. Thus we assume that *γ* grows monotonically with the number of ‘tests per case’. Another measure related to ‘tests per case’ and *γ* is the ratio of the delayed daily new cases to daily new deaths. A large number of ‘cases per death’ indicates that a large fraction of infected individuals have been identified (and isolated), and thus implies large *γ*.

In the third phase, while mobility is increasing, *R* is smaller than 1 and remains nearly constant, suggesting that the inevitable growth of *β* is compensated by a proportionate growth of *γ*. Indeed, when *R* is kept below 1, there is epidemic regression, during which the number of daily new cases is steadily decreasing. This leads to an increase of the average number of tests per case as can be seen for nearly all countries in Fig. 3. As a consequence, a given value of mobility reduction (and, thus, of *β*) corresponds to two values of *R*: higher than 1 in the second phase (when *γ* is low) and lower than 1 in the third phase (when *γ* is high).

Considered countries that introduced restrictions when the number of daily cases was relatively small were able to perform a higher number of tests per case and thus achieved higher *γ*. This allowed them to reduce *R* below 1 at larger *β* values, that is, at smaller mobility reduction (see Fig. 2). This explains a positive correlation between the cumulative death toll and cumulative lockdown (as shown in Fig. 1c), implying that, when the epidemic eradication or suppression is the objective, there is no trade-off between public health costs and economic loss.

For most locations, data on COVID-19-registered deaths as well as lockdown level become available with a short delay. We thus consider these values as rapid indicators of life/health costs and of economical/social costs. At the time of manuscript revision it has become already possible to juxtapose the GDP decrease in the first half of 2020 with cumulative lockdown in this period. As one can see in Fig. 4, these two quantities are correlated with Pearson’s correlation coefficients *ρ* = 0.81 and *ρ*_w_ = 0.81.

**Figure 4:**
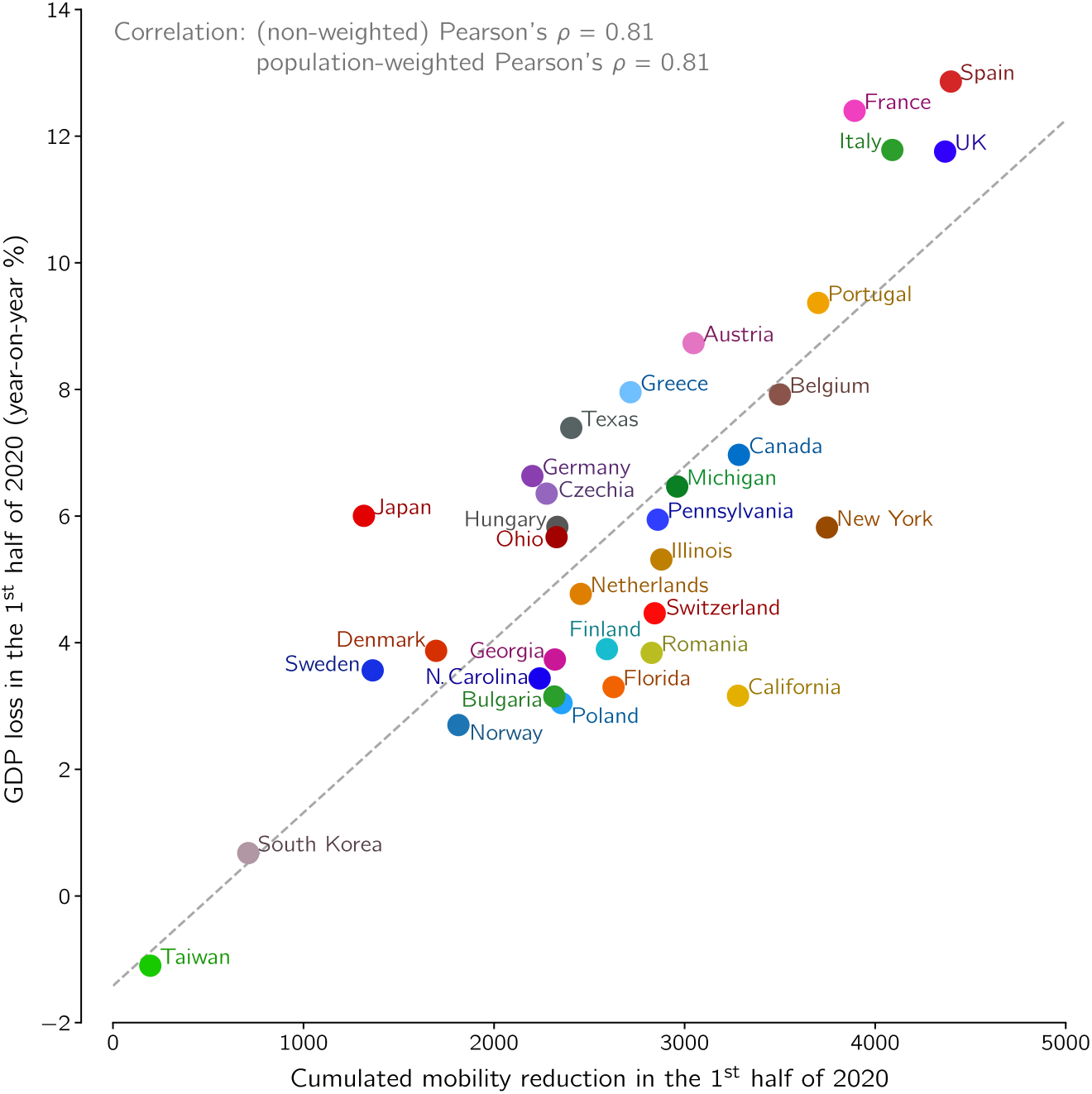
GDP loss in the first half of 2020 *vs*. cumulative lockdown in this period for European and East Asian countries, ten US states, and Canada.

Finally, we use the recently available data and directly juxtapose the population-normalized number of epidemic-associated deaths in the first half of 2020 with the GDP decrease (on a year-on-year basis) in this period of time, see Fig. 5. The epidemic-associated deaths are here defined as a maximum of the number of COVID-19-assigned deaths and excess deaths (the difference between actual all recorded deaths and their expected value based on recent years [6, 7]). We propose to take the maximum of these two values as for a handful of countries the death excess in the first half of 2020 is negligible (or even negative) due to the preventive lockdown which also reduced deaths associated with, e.g., car accidents, influenza, or air pollution [8, 9]. In the first wave of the epidemic in some considered countries, such as Spain or Netherlands, the difference between COVID-19-assigned deaths and excess deaths was significant (about 40% of excess deaths), whereas in others, such as France or Belgium, it was negligible [10]). The data from the second wave suggest that in some Central European countries (e.g., Poland and Bulgaria) the difference between the number of COVID-19-assigned deaths and excess deaths is larger than 40% of the latter. Obviously, the excess deaths (that are counted with some delay) serve as a better indicator of health-related damage than COVID-19-assigned deaths. When all countries and states are considered jointly, as in Fig. 4, correlation between the square root of the population-normalized number of epidemic-associated deaths and GDP is *ρ* = 0.55 and *ρ*_w_ = 0.68. When only-European countries are considered, correlation rises to *ρ* = 0.68 and *ρ*_w_ = 0.81, however this subset of data does not fulfill normality assumption according to majority of tests (see Methods for details) and thus should be considered with caution.

**Figure 5:**
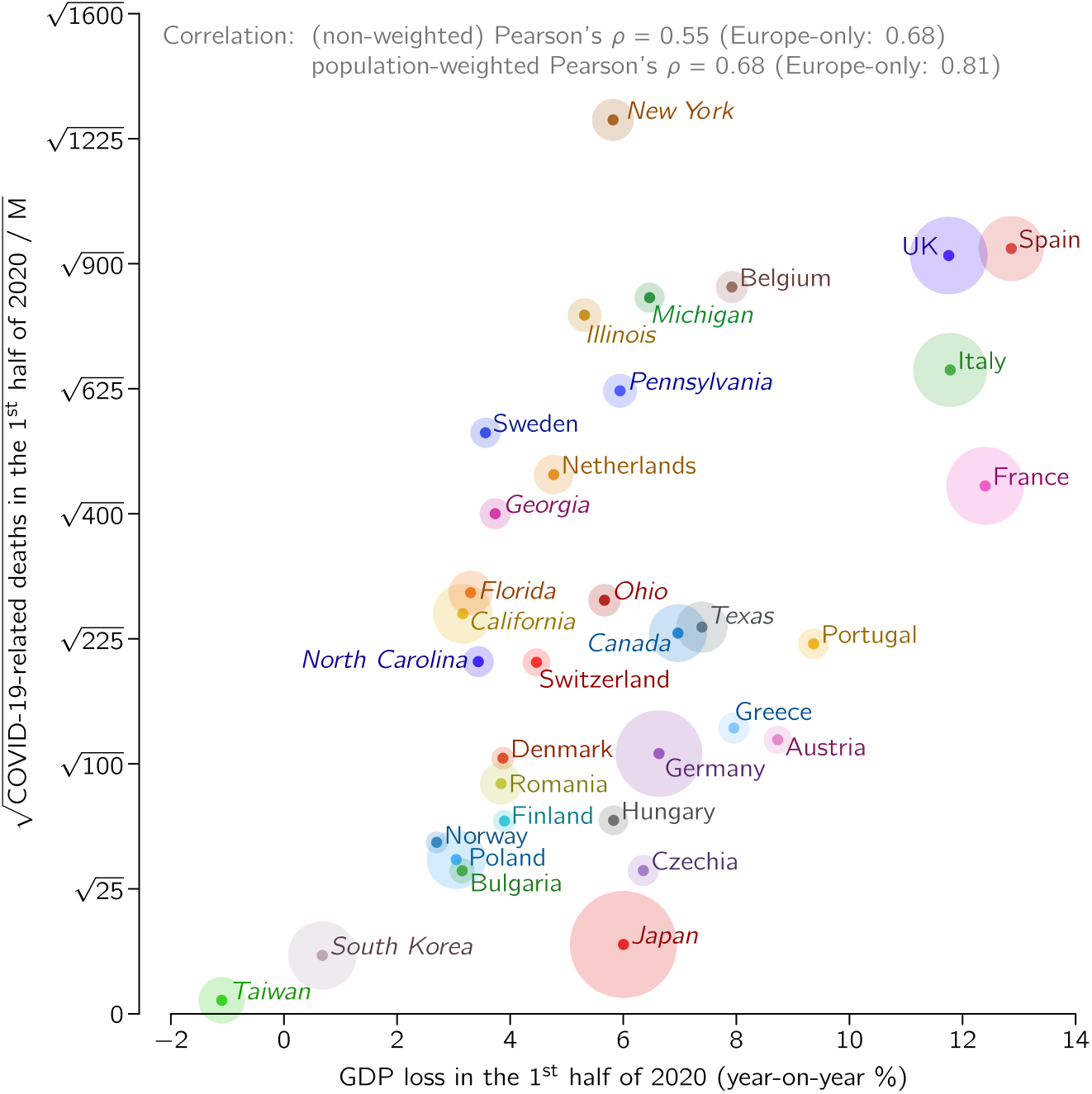
GDP loss in the first half of 2020 *vs*. square root of population-normalized number of epidemic-associated deaths in the same period for European and East Asian countries, ten US states, and Canada. Disk area of each location is proportional to its population.

The difference is likely caused by different socio-economic conditions in the US, Europe, and Japan. Gini index [11] for the US is 0.39, which is higher than for EU, 0.31, and Japan, 0.34, and is associated with lower social security level. Consequently, in the US the pandemic caused a sharp unemployment surge of 10 percentage points, while in the EU and Japan the rise of unemployment was close to 1 percentage point (first half of 2020 with respect to the first half of 2019) [12–14]. Thus, in the EU and Japan the GDP drop is the main consequence of lockdown, while in the US a comparatively smaller drop of GDP is accompanied by a higher increase of unemployment. Nevertheless, even when restricting to a single economical indicator, GDP, the correlation observed in Fig. 5 shows that during pandemic saving lives and protecting economy are not the opposite objectives.

## Discussion

Construction of Pareto fronts allowed us to identify countries that, up to a given time point, minimized the socio-economic and medical costs more effectively than others. Economic and social costs associated with lockdown include GDP loss, surge of unemployment, as well as educational and psychological costs caused by closing of schools and universities. Analysis of the latter costs requires additional studies as countries significantly differ in the stringency of restrictions imposed on their educational institutions. Medical costs are deaths but also severe cases, number of which is proportional to the number of deaths. Severe cases may be associated with lasting health effects. Additionally, high number of COVID-19 cases impact healthcare systems, aggravating treatment of other conditions.

Without weights being assigned to cumulative deaths and to the cumulative lockdown, one may not compare countries in a given Pareto front. Nevertheless, constructed layered Pareto fronts clearly show that the considered East Asian countries outperformed European countries and US states in both categories. These East Asian countries are on the “good” side of the European Pareto-optimal front formed by Slovakia, Norway, Greece, and Denmark (as of November 1, 2020). The Pareto-pessimal front is formed by UK (with the highest cumulative lockdown), Belgium, and New York State (with the highest cumulative population-normalized number of deaths). These locations, as well as locations in the second Pareto-pessimal front, at first suffered from a high number of deaths due to the delayed lockdown decision and then had a long and stringent lockdown introduced to stop the spread of the epidemic. Short Pareto fronts that clearly separate countries and states that up to a given time point perform better than others are the consequence of the strong correlation between (square root of) the population-normalized number of cumulative deaths and cumulative lockdown.

We have explained this non-intuitive correlation by analysis of trajectories of individual countries and states on the mobility reduction *vs*. effective reproduction number plane. We have shown that when the number of daily new cases exceeds some threshold, further epidemic growth can be suppressed only by a strict quarantine. In the growth phase, the number of tests per case decreases, implying reduction of the removal rate *γ*. This in turn implies that the reproduction number *R* = *β*/*γ* is decreased only due to the lockdown that reduced the contact rate *β*. Only after the imposed quarantine reduced *R* below 1 can the epidemic be further suppressed by the increased efficiency of testing (number of test per case), increasing the removal rate *γ*. The increase of *γ* allows for curbing the number of daily new cases (and resulting deaths) at gradual relaxation of lockdown measures.

To recognize reasons of vast differences in the number of cumulative deaths between countries after the first wave of the epidemic one can consider two hypothetically identical countries. Let us assume that in both countries in each week the number of daily cases grows twofold (which roughly corresponds to *R* = 2). Then, the first country decides to impose a strong lockdown which eventually (with some time delay) causes that the number of daily cases decreases 1.4-fold each week (which roughly corresponds to *R* = 0.7, which is the value achieved by Italy or Spain). If the second country introduces lockdown of the same stringency but delayed by one week, it will reach a twofold higher number of daily cases and additionally will have to sustain its lockdown 2 weeks longer in order to reduce the number of daily cases to the same level as the first country. Having twofold higher peak of daily cases and three-week longer epidemic wave, the second country will suffer from more than doubled count of deaths with respect to the first country and additionally will have higher economic loss due to the longer lockdown.

Over time, countries may change their Pareto fronts. The “velocity” of a country in the cumulative lockdown *vs*. cumulative deaths plane is given by the number of daily deaths and current mobility reduction. In a bit longer perspective, countries having currently *R* > 1 are in the unfavorable situation as their number of daily new cases is still increasing. In a longer time scale, both the cumulative death toll and the country-specific history of lockdown (lockdown fatigue) should be taken into consideration when predicting plausible further country trajectory. Clearly, these two factors influence risk avoidance and risk acceptance, respectively. Societies that managed to prevent high death toll by early and effective lockdown may find themselves in a psychologically more challenging situation as they will face lockdown fatigue that will not be counterbalanced be the “accumulated fear”. On the other hand, in countries that had a high wave of deaths, the accumulated fear is overshadowed by accumulated lockdown fatigue. In these countries the lockdown was more stringent and longer compared to countries that avoided the first wave of the epidemic by preventive lockdown. Comparison of the current situation (as of November 2020) in Central European countries that avoided the first wave of epidemic (due to preventive lockdown) with the situation in Western European countries that suffered from both a high death toll and long and stringent lockdown does not give a clear answer about their preparedness to the second wave. Currently, both Western and Central European countries, as well as the US, are attempting to contain the second wave without imposing lockdowns as stringent as during the first epidemic wave. Our analysis, however, suggests that this strategy can again lead to a high death toll and high economic and social costs.

An important assumption in our reasoning is that all analyzed countries continue the suppression strategy until the epidemic is terminated by a widespread immunization through vaccination. The relation between health and economic costs can be different when countries abandon the suppression strategy and chose the herd immunity option, in which the administratively imposed restrictions are replaced by voluntary risk-avoiding and risk-adapting strategies of the individuals [5]. In this case the preceding transient lockdowns may be not beneficial as they add economic costs to the high and unavoidable death toll.

## Conclusions

We have demonstrated that in the epidemic mitigation strategy, non-intuitively, there is no trade-off between the (population-normalized) number of deaths and the cumulative lockdown: these two variables are strongly correlated. Furthermore, the cumulative lockdown correlates with GDP decrease. Countries or states that refrained from imposing lockdown before the surge of infections suffered form both high numbers of epidemic-associated deaths as well as delayed but stringent and long-lasting lockdown. The “hit hard, hit fast” strategy, although difficult to implement for political reasons, appears to be the least devastating option. Strategies of individual countries (or states) can be evaluated with the help of Pareto fronts that stratify groups of countries with respect to the success of their anti-epidemic policies, without weighting medical and economic costs. The ordered ladder of short Pareto fronts clearly indicates that saving lives and protecting economy are not the opposite objectives.

## Methods

### Data sources

The COVID-19 dataset maintained by Our World in Data [15] is used as the source of data on the number of confirmed cases and deaths (original data source being European Centre for Disease Prevention and Control) and the number of tests (collected from national government reports). For the states of the USA, analogous data were extracted from The COVID Tracking Project [16]. Mobility reduction has been taken from the Google COVID-19 Community Mobility Reports [2].

Missing counts of daily new cases and deaths are filled with zeros and missing cumulative counts of cases and deaths are taken from the previous available counts. When the number of tests is not available daily, their number is linearly interpolated between available data points. To avoid artifacts in analyzed trends that would result from temporally non-uniform reporting, negative daily new counts were replaced by zeros while positive daily new counts known to result from aggregation of counts from multiple days were replaced by an average of counts in adjacent days.

Testing density [17] is analyzed according to: the number of people tested: in Canada, Netherlands, Norway, South Korea, Sweden; the number of samples tested: in Finland, Poland and Portugal; and the number of tests performed: for all remaining countries; for Belgium and Ireland units are labeled as unclear.

### Estimation of the reproduction number, *R*

To compute *R* based on the daily new confirmed cases (Fig. 2) we first estimated the doubling time *T*_d_ at each *i*th day as:

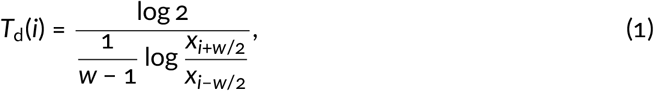

where *x*_*i*_ is rolling mean of the respective daily new counts (calculated within a 14-day window with equal weights) and the time span is set to *w* = 14 days. Let us notice that *T*_d_ is positive when the number of daily cases (or deaths) is increasing, and *T*_d_ is negative when the number of daily cases is decreasing; when the number of daily cases remains constant in time, then *T*_d_ = ∞. The reproduction number *R* on *i*th day was then computed in accordance with Wallinga & Lipsitch (2007) Wallinga and Lipsitch [18] and Wearing *et al*. (2005) Wearing, Rohani, and Keeling [19] as:

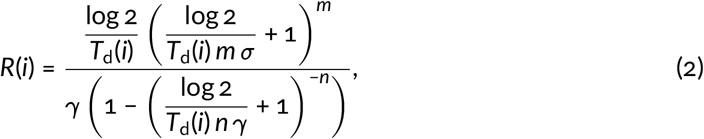

assuming *m* = 6, *n* = 1, *σ* = 1/(5.28 days), *γ* = 1/(2.9 days). These parameter values reflect the assumption that the incubation period is Erlang-distributed with the shape parameter 6 and mean of 5.28 days [20], while the period of infectiousness is distributed exponentially with the mean of 2.9 days. As a simplification, to estimate *R* the identical and constant in time *γ* has been assumed in all countries, however, as discussed in the Results section, the period of infectiousness is affected by testing capacity. For given *m, n*, and *σ* the reproduction number *R* is a decreasing function of *γ* for *T*_d_ > 0 and an increasing function of *γ* for *T*_d_ < 0. For *T*_d_ = ∞, we obtain *R* = 1, regardless of remaining parameters; thus the critical point in which *R* passes 1 in Fig. 2, is independent of the specific values of model parameters.

### Quantification of mobility reduction

To account for mobility reduction related to the decrease of economic activity, we averaged daily mobility reduction in categories of ‘workplaces’ and ‘retail and recreation’ (includes shopping centers and restaurants) on weekdays, excluding days of country-wide holidays in each country. National epidemic trajectories shown in Fig. 2 are drawn by taking current *R*(*i*) (computed based on the daily new cases as described above) and mobility reduction in previous days weighted according to the (time-reversed) distribution of infection-to-removal times, which is the hypoexponential distribution with rates {*λ, λ, λ, λ, λ, λ, γ*} where *λ* = 6*σ*.

### Normality of distributions

We confirmed normality of marginal distributions of points, for which Pearson’s correlation coefficient was computed, using an array of statistical tests as shown in Table 1.

**Table 1:**
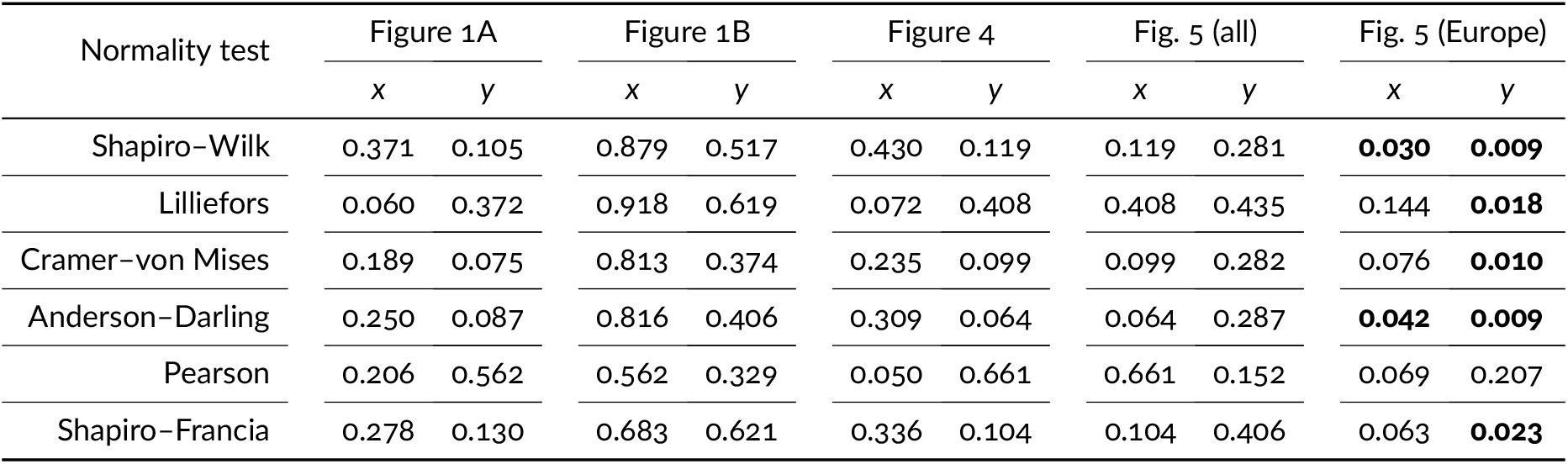
*P*-values from normality tests performed for data for which Pearson’s correlation coefficient is estimated. Columns labeled *x* and *y* correspond to projections of data points onto the horizontal and the vertical axis (marginal distributions), respectively. Values of *p* < 0.05 (bold) indicate violation of the normality assumption.

## Data Availability

All data are available within the article.

https://github.com/kochanczyk/covid19-pareto/

## Acknowledgments

This study was supported by the National Science Centre (Poland) grant 2018/29/B/NZ2/00668.

## Author contributions

M.K. conceived and performed the study and wrote the manuscript. T.L. conceived the study and wrote the manuscript.

## Corresponding authors

Correspondence to Marek Kochańczyk or Tomasz Lipniacki.

## Data availability

All data used in this study are referenced. Source code to analyze data and generate figures has been deposited at https://github.com/kochanczyk/covid19-pareto.

## Supplementary Information

**Supplementary Figure S1:**
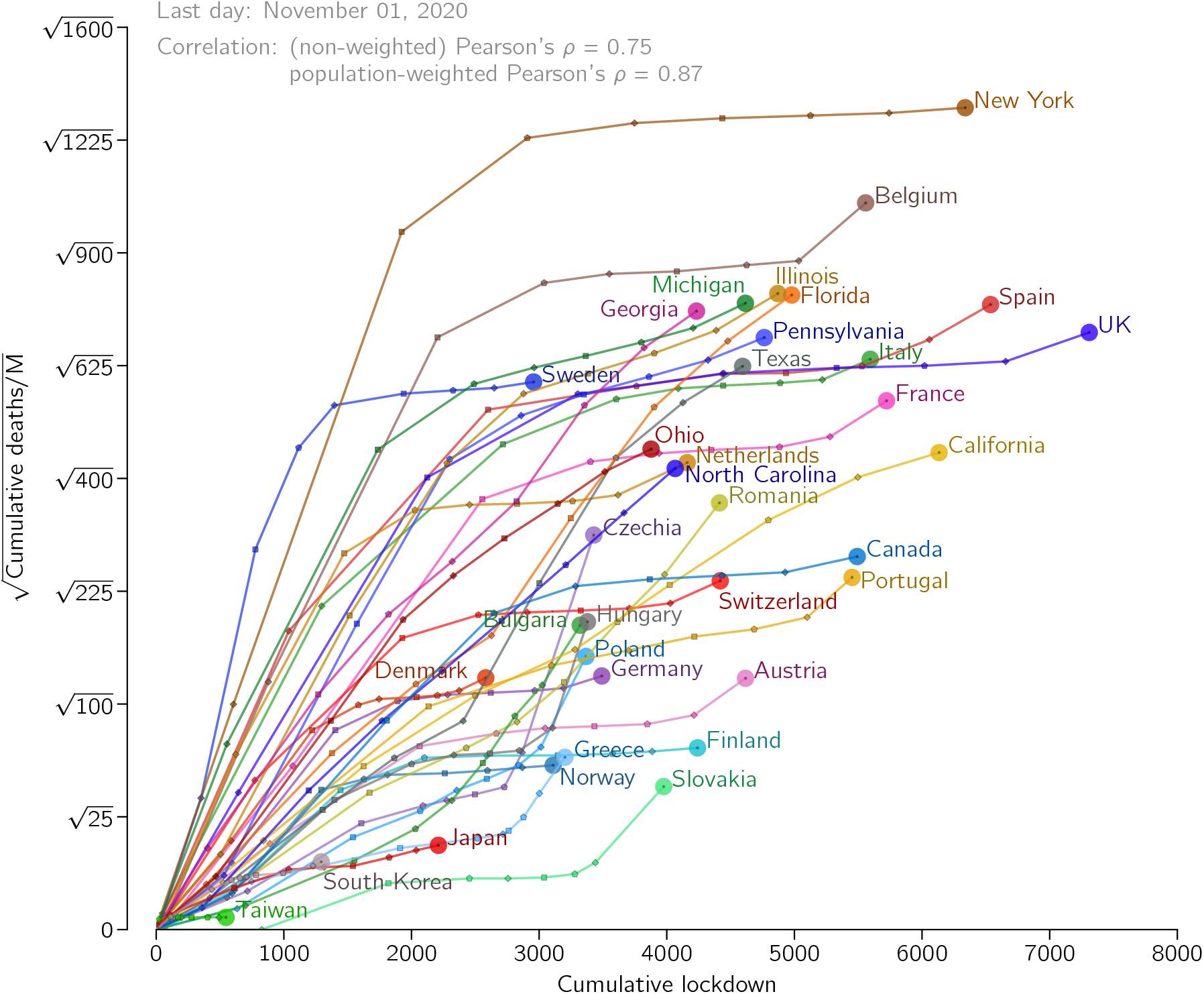
Time trajectories of countries and US states analyzed in Fig. 1 in the main text in the cumulative lockdown vs. square root of the population-normalized cumulative number of COVID-19-assigned deaths plane. Trajectories begin in February and end on November 1, 2020, and have a one-month resolution.

## References

[1] Roy M. Anderson and Robert M. May. Infectious diseases of humans: Dynamics and control. New York: Oxford University Press, 1991.

[2] Google LLC. Google COVID-19 Community Mobility Reports. https://www.google.com/covid19/mobility. accessed: 2020-11-09.

[3] Our World in Data. Human Development Index (HDI). https://ourworldindata.org/human-development-index. accessed: 2020-11-12.

[4] Zhongwei Huang et al. “Optimal temperature zone for the dispersal of COVID-19.” In:Science of The Total Environment 736 (2020), p. 139487. ISSN: 0048-9697. DOI: https://doi.org/10.1016/j.scitotenv.2020.139487.

[5] Marek Kochańczyk, Frederic Grabowski, and Tomasz Lipniacki. “Dynamics of COVID-19 pandemic at constant and time-dependent contact rates.” In: Mathematical Modeling of Natural Phenomena 15 (2020), p. 28. DOI: https://doi.org/10.1051/mmnp/2020011.

[6] Eurostat. Deaths by week and sex. https://ec.europa.eu/eurostat/databrowser/view/demo_r_mwk_ts. accessed: 2020-11-12.

[7] CDC National Center for Health Statistics. Excess deaths associated with COVID-19. https://www.cdc.gov/nchs/nvss/vsrr/covid19/excess_deaths.htm. acessed: 2020-11-12.

[8] Vasilis Kontis et al. “Magnitude, demographics and dynamics of the effect of the first wave of the COVID-19 pandemic on all-cause mortality in 21 industrialized countries.” In: Nature Medicine (2020). ISSN: 1546-170X. DOI: https://doi.org/10.1038/s41591-020-1112-0.

[9] Xinbo Lian et al. “Environmental indicator for effective control of COVID-19 spreading.” In: medRxiv (2020). DOI: https://doi.org/10.1101/2020.05.12.20099804.

[10] Our World in Data. A pandemic primer on excess mortality statistics and their comparability across countries. https://ourworldindata.org/covid-excess-mortality. acessed: 2020-11-12.

[11] OECD. Income inequality (indicator). https://dx.doi.org//10.1787/459aa7f1-en. accessed: 2020-11-12.

[12] Bureau of Labor Statistics. The employment situation. https://www.bls.gov/news.release/pdf/empsit.pdf. accessed: 2020-11-28.

[13] Eurostat. Unemployment by sex and age. https://ec.europa.eu/eurostat/databrowser/view/une_rt_m. accessed: 2020-11-12.

[14] Statistics Bureau of Japan. Labour Force Survey. Error! Hyperlink reference not valid.. accessed: 2020-11-28.

[15] Max Roser et al. Coronavirus Pandemic (COVID-19). https://ourworldindata.orgcoronavirus. accessed: 2020-11-09. 2020.

[16] The COVID Tracking Project. https://covidtracking.com. Accessed: 2020-11-09. 2020.

[17] Joe Hasell et al. “A cross-country database of COVID-19 testing.” In: Scientific Data 7.1 (2020), p. 345. ISSN: 2052-4463. DOI: https://doi.org/10.1038/s41597-020-00688-8. xURL: https://doi.org/10.1038/s41597-020-00688-8.

[18] Jacco Wallinga and Marc Lipsitch. “How generation intervals shape the relationship between growth rates and reproductive numbers.” In: Proceedings of the Royal Society B 274 (2007), pp. 599λ604. DOI: https://doi.org/10.1098/rspb.2006.3754.

[19] Helen J Wearing, Pejman Rohani, and Matt J Keeling. “Appropriate Models for the Management of Infectious Diseases.” In: PLOS Medicine 2 (2005), p. 0020174. DOI: https://doi.org/10.1371/journal.pmed.0020174.

[20] Stephen A. Lauer et al. “The Incubation Period of Coronavirus Disease 2019 (COVID-19) From Publicly Reported Confirmed Cases: Estimation and Application.”In: Annals of Internal Medicine 172.9 (2020), pp. 577λ582. DOI: https://doi.org/10.7326/M20-0504.

